# AI-Based Screening Approach for Primary Aldosteronism Using Longitudinal Electronic Health Records

**DOI:** 10.64898/2026.10.06.26364909

**Authors:** Frank G Lee, Ashok Choudhary, Leo Y. Li-Han, Chris Varghese, Ellen L Larson, William RG Perry, Irina Bancos, Trenton R Foster, Travis J McKenzie, Hojjat Salehinejad, Cornelius A Thiels

## Abstract

**Context:** Current guidelines for primary aldosteronism (PA) recommend screening all patients with hypertension. Given low screening rates, an automated artificial intelligence (AI)-based risk-stratified approach may facilitate guideline implementation.

**Objective:** To develop an AI-based prescreening tool using electronic health record (EHR) data to identify at-risk patients.

**Design:** Model development retrospective cohort study. Ensemble architecture using eXtreme Gradient Boosting, random forest, and extremely randomized trees was trained on EHR data (1986-2025) and evaluated on a test set of patients with hypertension.

**Setting:** Single institution system spanning community clinics and tertiary/quaternary centers reported via Mayo Clinic Platform.

**Patients:** Adults with hypertension, International Classification of Disease (ICD) diagnosis of PA, or had plasma renin/aldosterone testing.

**Main Outcome Measures:** Predicting PA diagnosis (disease) or negative workup (control) using age, sex, ICD diagnoses, vitals, labs, and medications by area under receiver operating characteristic (AUROC), estimated calibration error (ECE), and specificity.

**Results:** Of 22,264 patients (1,833 disease and 20,431 controls), median time from initial encounter to screening was 5.0 years (IQR 0.5-12.6). The model demonstrated AUROC 0.709 (95% CI 0.688-0.730) with ECE 0.037. At a 0.5 cut-off threshold, the model had high specificity (71.6%) and low false positivity (28.4%). In a test cohort of 1,279,455 adults with hypertension, the model identified 5,295 (0.4%) for first-pass screening.

**Conclusions:** A high-specificity model can provide a pre-screening PA pathway using routine EHR data to identify and prioritize patients for PA screening. This AI-based approach can risk-stratify patients, guide sequencing of testing/referrals, and pragmatically facilitate adoption of current guidelines into practice.

## Introduction

Approximately 129 million adults in the United States have hypertension (1,2). Primary aldosteronism (PA) is a leading cause of secondary hypertension with estimated prevalence up to 14%, however less than 2% of patients with hypertension are screened (3,4). Missed or delayed PA diagnosis has significant ramifications on patients and healthcare costs, resulting in higher rates of long-term cardiovascular/renal complications (5). Effective therapies exist for PA including mineralocorticoid receptor antagonists (MRA) with emerging medical options like aldosterone synthase inhibitors (ASI) increasingly recognized (6,7). In patients with unilateral PA, adrenalectomy can be curative however this opportunity is missed in patients without accurate diagnosis of PA (8).

To address this gap, current guidelines recommend screening all adults with hypertension (5). However, adoption has lagged as the resulting increase in testing may overwhelm current resource capabilities and potentially lead to healthcare overutilization, over-testing, and low-value care (9). Targeted approaches may improve pretest probability of positive PA screening by pragmatically improving the yield of positive case referrals to endocrinology. An artificial intelligence (AI) prescreening strategy applied at the population level could substantially close the gap. AI could address current limitations by identifying at-risk patients earlier, risk-stratify patients for testing, and limit false negative referrals to endocrinology in a way that traditional risk calculators cannot. By exploiting routine information available in the electronic health record (EHR) like International Classification of Disease (ICD) diagnosis codes, laboratory values, and medications, machine learning (ML) models can provide a potential viable prescreening strategy (11–14).

The aim of this study is to develop a population-level prescreening tool for PA that analyzes longitudinal EHR data. We hypothesize that an ML approach can discriminate patients for PA screening with 1 year lead-time. The resulting model can augment current guidelines to screen all patients with hypertension by risk-stratifying and sequencing the population in a targeted fashion. Lastly, we assessed the feasibility of our model by applying it to a large real-world cohort of adults with hypertension.

## Materials and Methods

This is a model development study design using retrospective data and is reported in accordance with TRIPOD+AI (15). Institutional policies and local laws were adhered to, which included Mayo Clinic Institutional Review Board approval (#25-010099).

### Data Source

This study used Mayo Clinic Platform, a HIPAA-compliant, multicenter database containing de-identified patient information regularly updated from participating institutions’ EHR systems using a federated, standardized data harmonization process (16). A data use agreement was signed on August 1, 2025 and is available in **Supplementary Material S1**. As of December 31, 2025, Mayo Clinic Platform contained over 8 million patient records from January 1, 1990 to December 31, 2025 from all Mayo Clinic sites including main campuses at Rochester, Minnesota; Jacksonville, Florida; and Scottsdale, Arizona; and smaller hospitals/clinics in the Mayo Clinic Health System (regional areas of Minnesota and Wisconsin). Access to Mayo Clinic Platform (https://mcpdiscover.org) was granted on August 26, 2025.

### Inclusion Criteria and Cohort Selection

Inclusion criteria were adults who met any of the following criteria: 1) had an ICD diagnosis of primary aldosteronism, 2) received a screening test i.e. plasma/serum aldosterone or plasma renin activity/concentration, or 3) had an ICD diagnosis related to hypertension (**Supplementary Material S2**). Patients with an ICD diagnosis of PA were assigned to the disease cohort in the training dataset; patients who received a screening test without a PA diagnosis were assigned to the control cohort in the training dataset; lastly, any remaining patients with a hypertension-related ICD diagnosis who had neither PA ICD diagnosis nor received a screening test were assigned to the test cohort (**Figure 1**).

**Figure 1:**
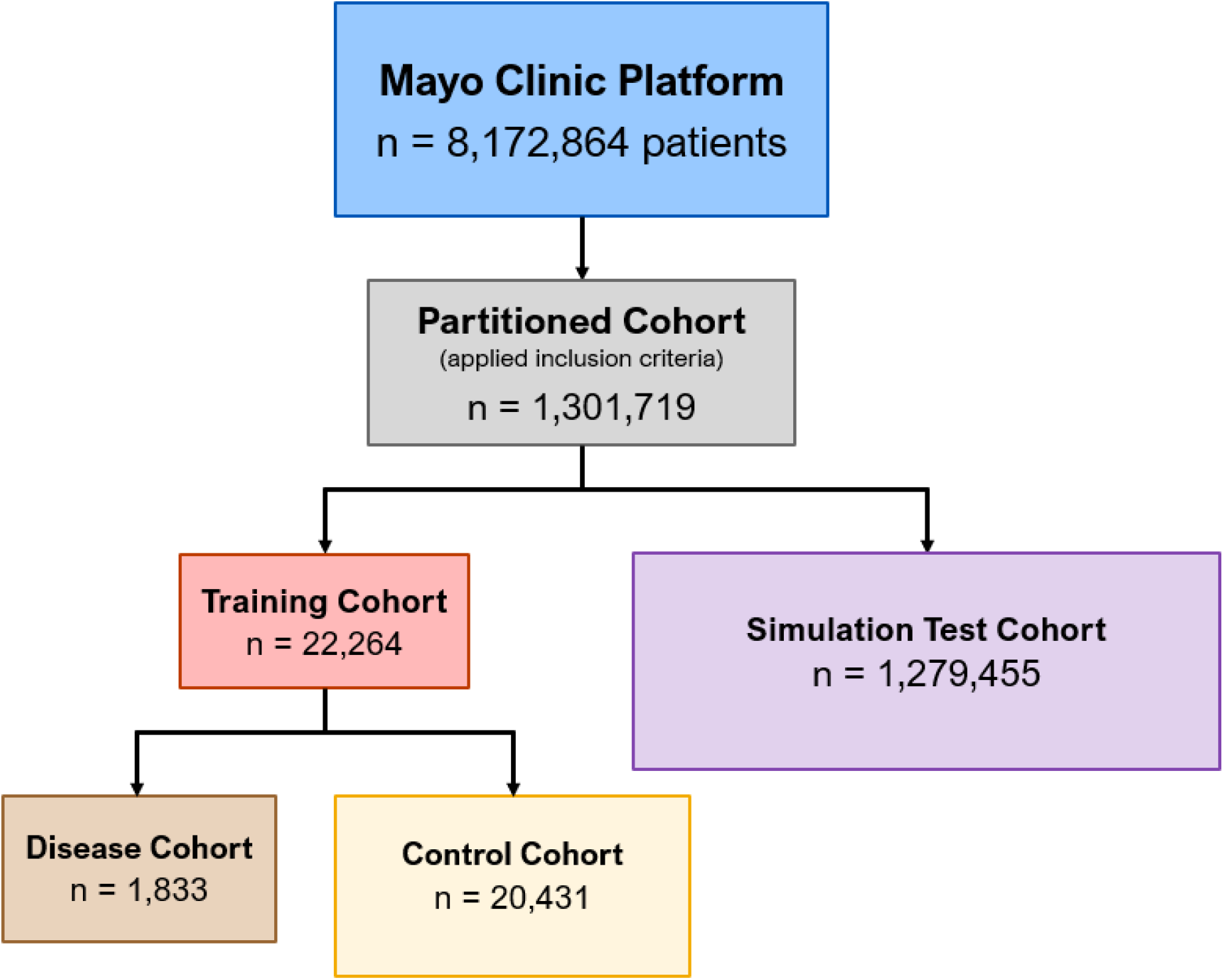
Cohort selection

### Reference Standard

Disease positive was defined as any patient who had an ICD-9/ICD-10 diagnosis code of primary aldosteronism: 255.12/E26.01 Conn’s syndrome, 255.12/E26.02 glucocorticoid-remediable aldosteronism, 255.10/E26.9 hyperaldosteronism, unspecified, E26.09 other primary hyperaldosteronism, and E26.89 other hyperaldosteronism. Disease negative was defined as any patient who had an occurrence of primary aldosteronism screening labs i.e. serum/plasma aldosterone and renin levels and never received an ICD diagnosis code of primary aldosteronism. This inferred approach of presence/absence of diagnosis codes and laboratory tests was determined to be the most feasible at the population level for the endpoint of timely screening. The use of biochemical laboratory cut-offs and pathology results were considered however implementation was limited due to the effects of confounders on equivocal results, i.e. potassium status and concurrent medications with inconsistent application and reporting. Principally relying on clinical diagnosis offered the most generalizability for model development. Patients with hypertension who never had serum/plasma aldosterone/renin testing were neither assigned to disease positive nor disease negative and therefore comprised the test cohort.

### Model Development

An ensemble model architecture was devised, which combined eXtreme Gradient Boosting (XGBoost), random forest, and extremely randomized trees (ExtraTrees) model. Model development and training occurred on Mayo Clinic Platform which provided computational resources and storage with on-demand virtual environments. Predictor variables for model training input were selected based on well-known features of PA: hypertension-related and hypokalemia-related ICD codes, systolic blood pressure (SBP) measurements, serum potassium levels, and oral prescription medications of antihypertensives and potassium supplementation (**Supplementary Material S4**) (17). Age and sex were also used as model inputs as age- and sex-specific differences exist (17,18). The dates and timestamps associated with predictor variables were collected for encoding longitudinal information. **Figure 2** illustrates the signal within a patient’s clinical history to be leveraged by the ML model.

**Figure 2:**
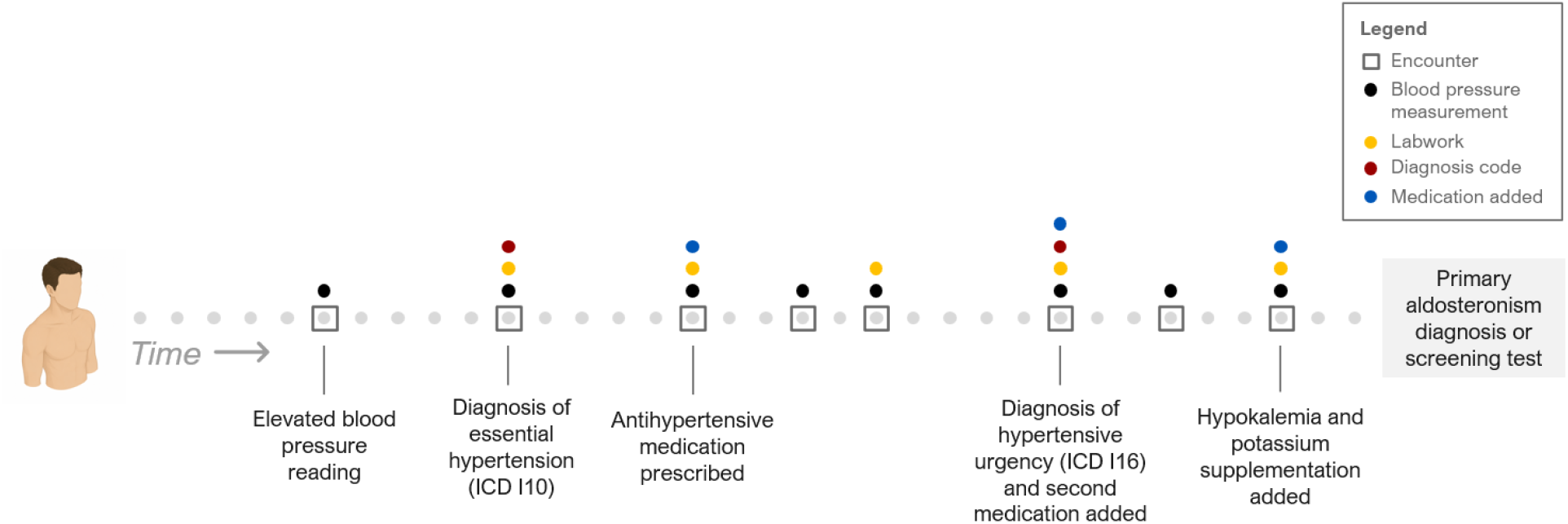
Illustration of representative patient clinical history showcasing timeline of input predictor variable

The model was trained to predict likelihood of PA diagnosis (disease) or negative PA testing (control). To incorporate early prediction capabilities and avoid potential recency testing bias immediately prior to diagnosis or screening outcome, a cut-off window (washout period) was implemented at 30-day and 1-year lead-times. We used t_0_ to denote time of outcome: date of PA diagnosis for the disease cohort or date of screening lab tests (i.e. serum aldosterone or plasma renin) for the control cohort. For 30-day lead-time, the model used only predictor variables with date/timestamps 30 days or more before t_0_. Similarly, for 1-year lead-time, only predictor variables with date/timestamps up to 1 year before t_0_ were included. Patients who lacked clinical histories of 30 days or 1 year before date of outcome were excluded from respective models. A zero was inputted into the tabular training data for cases where a predictor variable category was not available, e.g. no associated ICD code or potassium lab was recorded in the patient’s EHR. The degree of missingness for each input predictor variable was reported directly in the results.

A hyperparameter search was performed for XGBoost using random sampling, and ultimately a maximum decision tree depth of 3, learning rate of 0.01 with 3200 boosting rounds (trees), subsample of 0.8, and L1 regularization (sparsity) of 100 was chosen for best performance. A similar search was used for random forest and ExtraTrees. More information is available in **Supplementary Material S5**. The model was internally developed using 5-fold cross-validation (i.e. an 80/20 training/validation split was employed in which 80% of patients were used for training and the remaining 20% reserved for internal validation and testing and repeated for a total of 5 folds, each with different partitions of training data). To address class imbalance, the disease cohort (minority class) was randomly upsampled 1:1 to match the controls (majority class). For validation, natural distribution (i.e. no correction of class imbalance) was used.

#### Model Performance and Feature Importance

The model’s predictive performance was assessed using area under the receiver operating characteristic (AUROC) curve and estimated calibration error (ECE). AUROC >0.7 was defined as acceptable (19). Secondary measures were threshold-dependent metrics and included sensitivity (recall), specificity, false negative rate, false positive rate, accuracy, precision (positive predictive value), and F1-score. Formula definitions are provided in **Supplementary Material S6**. Calibration plots were prepared for both 30-day and 1-year predictions. Metrics were calculated for each fold, and the mean was reported with 95% confidence interval (CI). Lastly, predictor variables were analyzed by feature importance to sort the contribution of these factors on predicting outcome for both 30-day and 1-year models.

#### Model Feasibility Testing

The trained model was applied to a retrospective test cohort of patients with hypertension who were never diagnosed with or screened for PA. The intent of this analysis is to simulate how the model would perform in a real-world environment. Different model thresholds (at decile intervals of 0.1) were queried to see how many patients would be identified for screening by the model. For this analysis, t_0_ was set to the latest predictor variable date i.e. pertinent ICD diagnosis, blood pressure, potassium lab, or medication related to hypertension or potassium supplementation. Both 30-day and 1-year models were applied to the test cohort with 30-day and 1-year training cut-off periods from t_0_, respectively. Lastly, a decision curve analysis (DCA) was performed to identify a clinically appropriate cut-off threshold for prescreening.

#### Statistical Analysis

Cohort characteristics were described with summary statistics using median and interquartile range (IQR) for continuous variables and count (n) and percentage (%) for categorical variables. Comparisons between disease and control cohort predictor variables were made using Wilcoxon rank sum (WR) and Chi-square (χ^2^) tests where p<0.05 was considered significant. Equity analysis was performed on patient self-reported race to ensure fairness and AUROC parity across racial subgroups of White/Caucasian, Black/African American, Asian, and other/unknown. Asian included Indian, and other/unknown included Native American and Pacific Islander, chose not to disclose, and multiple races. Analysis was performed using Python version 3.12 (Python Software Foundation, Wilmington, Delaware).

## Results

A total of 22,264 patients (1,833 disease and 20,431 control) were identified for training with median age 66 (IQR 52-76), 12,039 (54.1%) female, and 19,831 (89.1%) White/Caucasian. Median follow-up from first model input predictor variable to outcome was 5.0 years (IQR 0.5-12.6). The disease cohort compared to control group had higher rates of hypertension- and hypokalemia-related diagnoses, higher SBP, lower potassium, and higher use of potassium supplementation medications (all p<0.001, **Table 1**). For model development, the level of missingness among predictor variables ranged from 27.0% for pertinent ICD codes to 70.9% for relevant medications (complete missingness information provided in **Supplemental Material S7**).

**Table 1:** Cohort characteristics for the disease and control cohort within the training dataset.

|  | **Disease Cohort**<br>N = 1,833 | **Control Cohort**<br>N = 20,431 | **p** |
| --- | --- | --- | --- |
| Age | 67 (55-76) | 66 (51-76) | <0.001<sup>w</sup> |
| Female | 862 (47.0%) | 11,177 (54.7%) | <0.001<sup>x</sup> |
| Clinical history, years | 4.0 (0.3-10.9) | 5.1 (0.6-12.9) | <0.001<sup>w</sup> |
| Hypertension-related ICD diagnosis | 1,407 (76.8%) | 14,545 (71.2%) | <0.001<sup>x</sup> |
| Hypokalemia ICD diagnosis | 285 (15.5%) | 1,819 (8.9%) | <0.001<sup>x</sup> |
| SBP reading | 132 (118-148) | 126 (111-142) | <0.001<sup>w</sup> |
| Potassium level | 3.9 (3.6-4.3) | 4.2 (3.9-4.6) | <0.001<sup>w</sup> |
| Number of antihypertensive medications | 2 (1-3) | 2 (1-3) | 0.433<sup>w</sup> |
| Receiving potassium supplementation | 314 (13.3%) | 1,857 (9.3%) | <0.001<sup>x</sup> |

Values reported as median (Q1-Q3) or N (%). w Wilcoxon rank-sum test, X Chi-square test

Discriminative performance of the model on predicting disease from negative screening was AUROC 0.769 (95% CI 0.749-0.789) for 30-day lead-time and AUROC 0.709 (95% CI 0.688-0.730) for 1-year lead-time (**Figure 3A**). ECE calibration values for 30-day and 1-year models were optimized to 0.036 and 0.037 and particularly well-calibrated for moderate risk probability ranges between 0.5-0.6. Threshold-dependent metrics are shown in **Table 2**. For a cut-off threshold of 0.5, the model had 60.0% sensitivity for 30-day and 57.2% sensitivity for 1-year. A 0.5 cut-off translated to a false positive rate of 21.1% for 30-day and 28.4% 1-year models (**Figure 4**). The model exhibited no bias in performance across racial subgroups comparing White/Caucasian, Black/African American, Asian, and other/unknown with AUROC 0.71-0.77 (p=0.58, **Supplementary Material S8**). Feature importance analysis showed mean potassium lab levels, days diagnosed with ICD-9 405.99 (other unspecified secondary hypertension), days diagnosed with ICD-10 E87.6 (hypokalemia), days on spironolactone and eplerenone medication contributed most to the model’s prediction (**Figure 5**).

**Figure 3:**
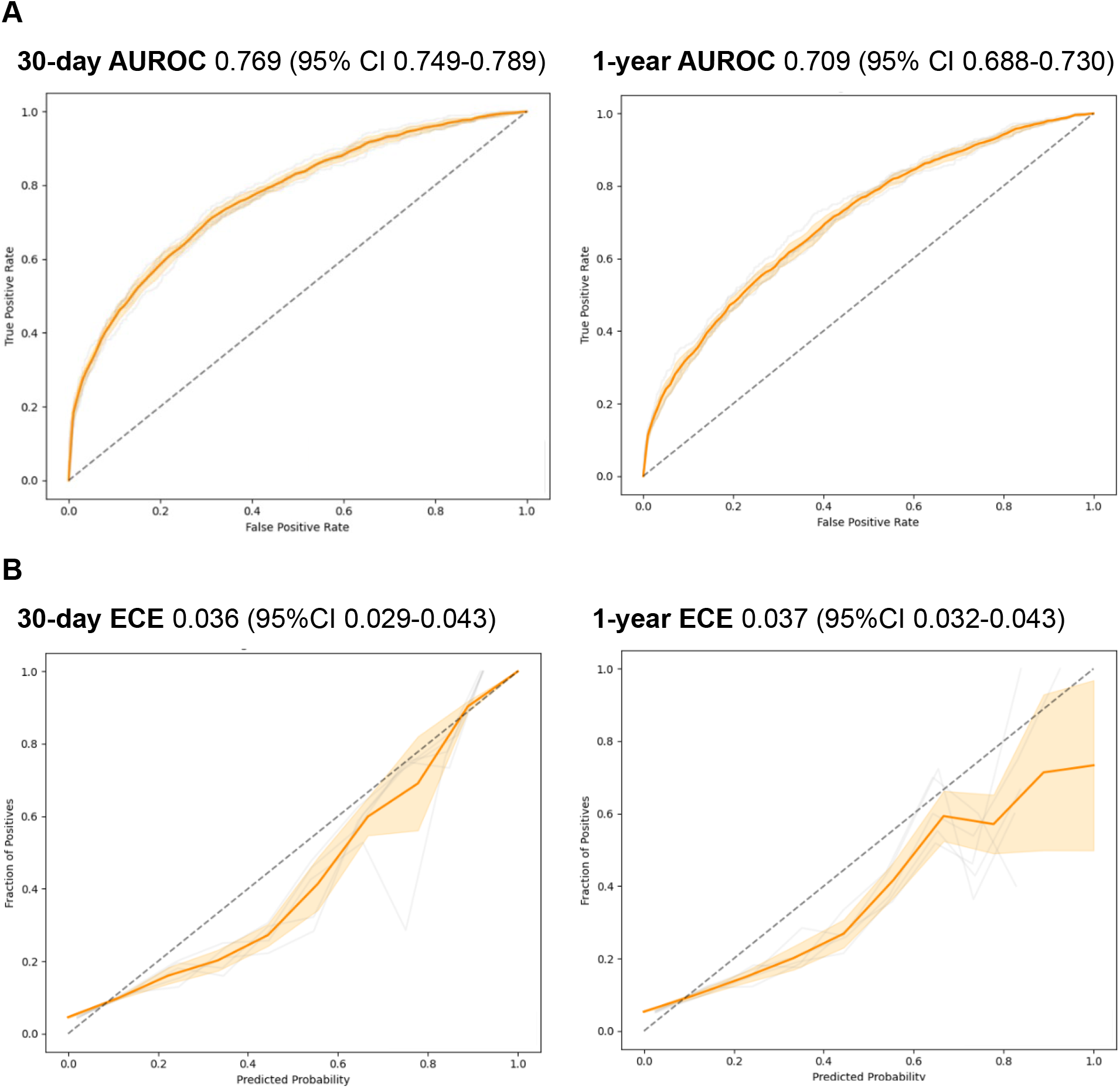
Results of 5-fold cross-validation from ensemble model development using (A) area under receiver operating curve (AUROC) discriminative performance and (B) calibration plots for 30-day and 1-year lead-times.

**Figure 4:**
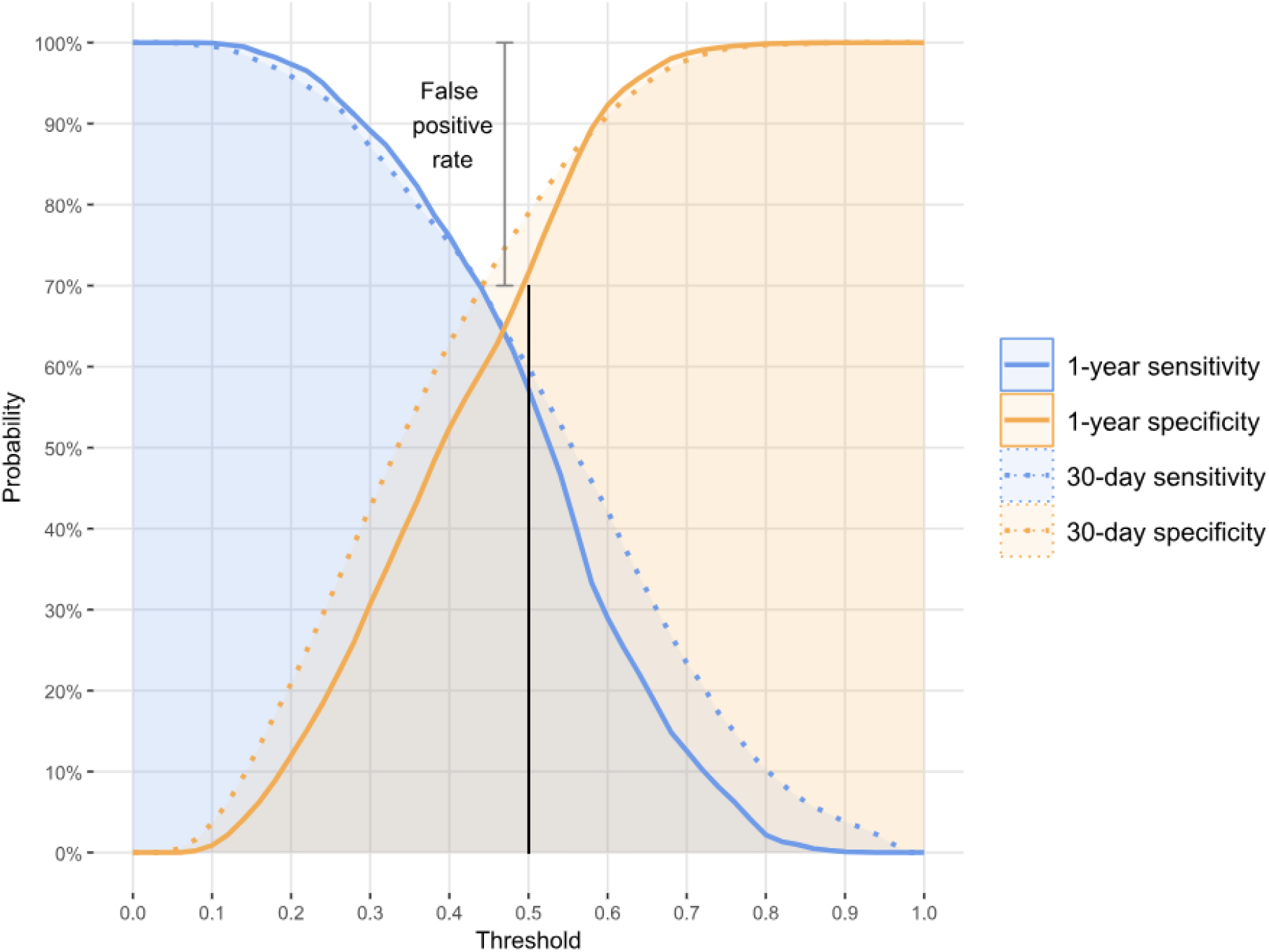
Sensitivity and specificity curves for the ensemble model with selected cut-off threshold of 0.5

**Figure 5:**
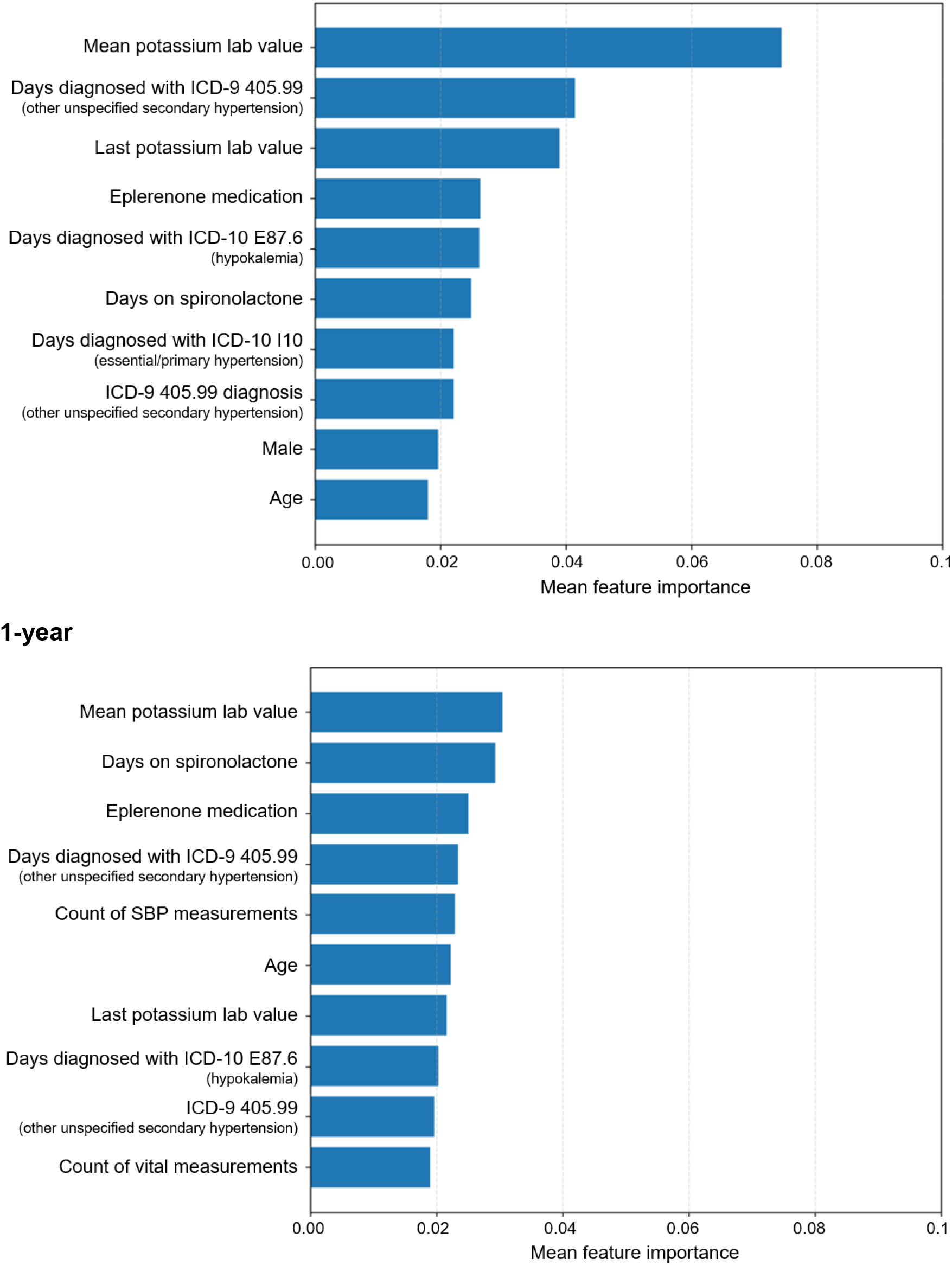
Feature importance analysis for model prediction (top 10 variables displayed)

**Table 2:** Sensitivity, specificity, accuracy, precision, and F1-score results of 5-fold cross-validation from ensemble model development for 30-day and 1-year lead-times at various cut-off thresholds.

| **30-day** |  |  |  |  |  |  |  |
| --- | --- | --- | --- | --- | --- | --- | --- |
| Threshold | Sensitivity<br>(Recall) | Specificity | False<br>negative<br>rate | False<br>positive<br>rate | Accuracy | Precision<br>(PPV) | F1-score |
| 0.1 | 0.996 | 0.036 | 0.004 | 0.964 | 0.115 | 0.085 | 0.156 |
| 0.2 | 0.959 | 0.209 | 0.041 | 0.791 | 0.271 | 0.098 | 0.178 |
| 0.3 | 0.871 | 0.426 | 0.129 | 0.574 | 0.463 | 0.120 | 0.211 |
| 0.4 | 0.753 | 0.629 | 0.247 | 0.371 | 0.639 | 0.154 | 0.256 |
| 0.5 | 0.600 | 0.789 | 0.400 | 0.211 | 0.773 | 0.203 | 0.304 |
| 0.6 | 0.422 | 0.909 | 0.578 | 0.091 | 0.869 | 0.295 | 0.347 |
| 0.7 | 0.234 | 0.978 | 0.766 | 0.022 | 0.917 | 0.490 | 0.317 |
| 0.8 | 0.103 | 0.997 | 0.897 | 0.003 | 0.924 | 0.770 | 0.181 |
| 0.9 | 0.038 | 1.000 | 0.962 | 0.000 | 0.921 | 0.972 | 0.072 |
| **1-year** |  |  |  |  |  |  |  |
| Threshold | Sensitivity<br>(Recall) | Specificity | False<br>negative<br>rate | False<br>positive<br>rate | Accuracy | Precision<br>(PPV) | F1-score |
| 0.1 | 0.999 | 0.009 | 0.001 | 0.991 | 0.090 | 0.083 | 0.153 |
| 0.2 | 0.973 | 0.120 | 0.027 | 0.880 | 0.190 | 0.090 | 0.165 |
| 0.3 | 0.891 | 0.307 | 0.109 | 0.693 | 0.355 | 0.103 | 0.185 |
| 0.4 | 0.761 | 0.524 | 0.239 | 0.476 | 0.543 | 0.125 | 0.215 |
| 0.5 | 0.572 | 0.716 | 0.428 | 0.284 | 0.704 | 0.153 | 0.242 |
| 0.6 | 0.290 | 0.923 | 0.710 | 0.077 | 0.871 | 0.252 | 0.269 |
| 0.7 | 0.125 | 0.986 | 0.875 | 0.014 | 0.916 | 0.454 | 0.197 |
| 0.8 | 0.022 | 0.999 | 0.978 | 0.001 | 0.918 | 0.581 | 0.042 |
| 0.9 | 0.001 | 1.000 | 0.999 | 0.000 | 0.918 | 0.200 | 0.002 |

PPV positive predictive value

When applied to a test cohort of 1,279,455 adults with hypertension, the model identified 5,295 (0.4%) for screening using a cut-off threshold of 0.5. Adjusting cut-off thresholds affected the total number of patients identified for screening by the model, providing a modifiability factor to AI-based screening methods (**Table 3**). Decision curve analysis supported the use of a high-specificity cut-off i.e. 0.5 for adding discriminating predictive performance compared to current screening standards of all patients with hypertension (**Figure 6**).

**Figure 6:**
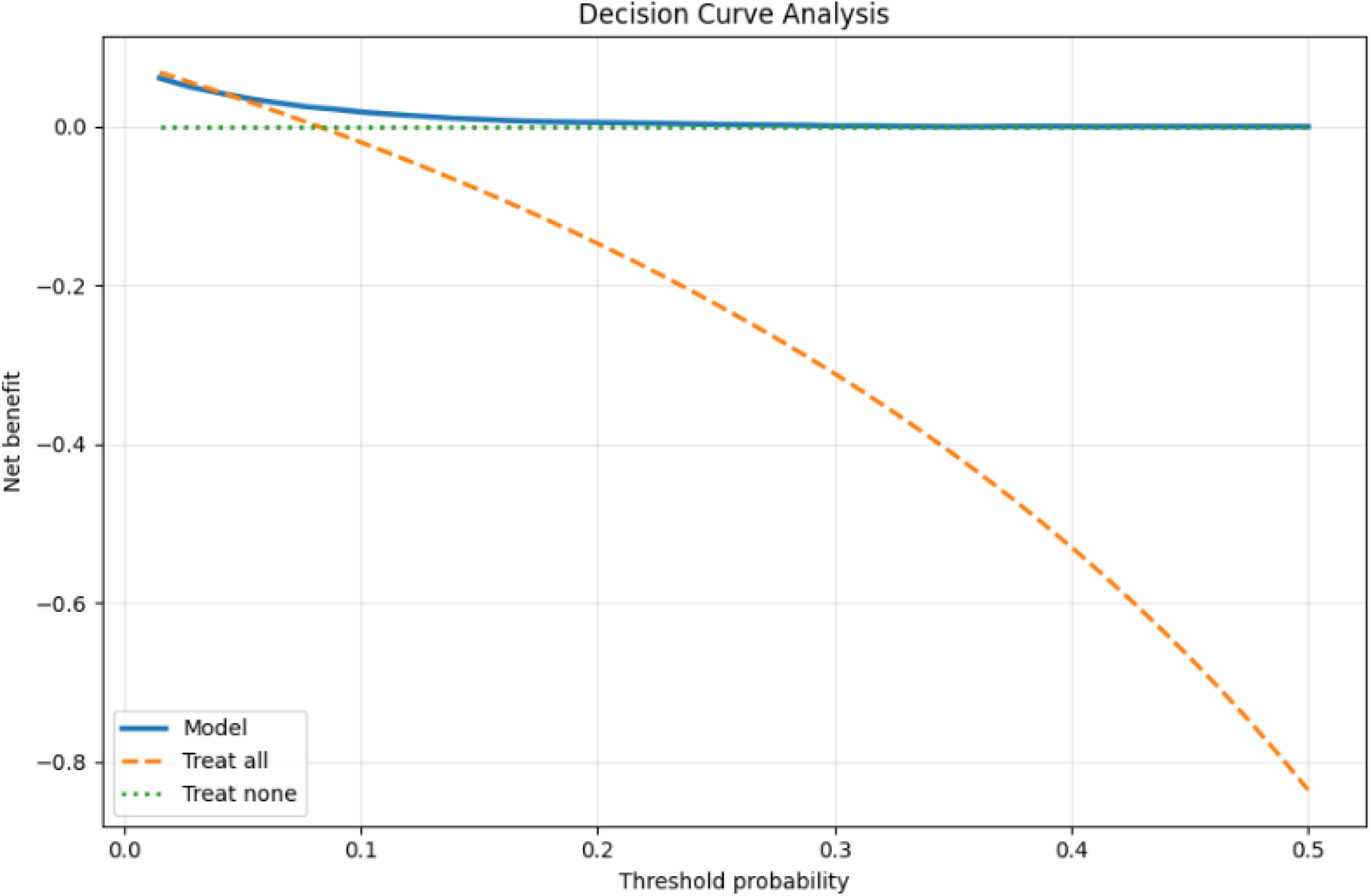
Decision curve analysis demonstrating that a selected cut-off threshold of XX yields the highest clinical net benefit compared to testing all patients with hypertension (orange dashed line) or no testing (green dashed line)

**Table 3:** Number of patients identified in test cohort of adults with hypertension (n=1,279,455) identified for screening by the ensemble model.

| Threshold | Number of patients identified for screening | Screening rate |
| --- | --- | --- |
| 0.1 | 1,279,200 | 99.98% |
| 0.2 | 1,163,150 | 90.91% |
| 0.3 | 706,680 | 55.23% |
| 0.4 | 96,560 | 7.55% |
| 0.5 | 5,295 | 0.41% |
| 0.6 | 637 | 0.05% |
| 0.7 | 77 | 0.006% |
| 0.8 | 21 | 0.002% |
| 0.9 | 0 | 0% |

## Discussion

An ML model trained on patients’ longitudinal EHR data achieved acceptable discriminative performance with well-calibrated risk estimates, supporting its use as a pragmatic tool to prioritize patients with hypertension for PA screening. This is a novel low-cost, automated, population-based strategy to effectively risk-stratify adults with hypertension for screening and serve as an adjunct to current guidelines. A cut-off threshold of 0.5 was an ideal setting for a prescreening approach to minimize false positive cases with good calibration. While current guidelines recommend universal screening for all patients with hypertension, this pragmatic approach may increase compliance and improve outcomes for patients. Decision curve analysis reinforced the notion that testing all patients diminishes net benefit and a high specificity prescreening model can yield a higher rate of true positives. Additionally, the ability to set specific thresholds is an ideal lever for institutions given resource constraints. Exploratory testing of the model on a large cohort of hypertensive adults without previous PA screening demonstrated feasibility and can provide a practical prescreening tool for healthcare systems to readily deploy and broadly implement due to the input of routinely available EHR data.

PA is underrecognized, under-screened, and underdiagnosed (20). Less than 10% of primary care providers have reported screening for PA (21). Large retrospective series show screening rates of less than 2% of patients (22), and delays in diagnosis are estimated to be a median of 4.5 years (23). The 2025 Endocrine Society guidelines attempt to address this by screening all patients with hypertension (Grade 2 Strength and Low Certainty of Evidence) (5). These recommendations align with other recent guidelines from the European Society of Cardiology and American Heart Association/American College of Cardiology, which recommend mass screening of patients with treatment-resistant hypertension or Grade 2 hypertension (≥140/90 mmHg) (4,24,25). However, practical steps towards implementing this at population-scale are lacking without solutions to address the increased demand and ensuing strain on resource capacity (26).

The Endocrine Society guidelines acknowledge that their recommendation is “conditional … with implementation depending on contextual factors such as available resources, local expertise, and healthcare system capacity, which may affect feasibility and prioritization” considering cost-effectiveness and new technologies for screening (5). We propose using AI as a practical and low-cost adjunct to ameliorate the demands on the healthcare system. The seminal work on principles of screening by Wilson and Jungner include *cost-effectiveness* and *ongoing endeavor* (27). A selective screening approach by AI may reduce over-testing and low-value care (diminishing quality with higher costs) when 47.7% of the U.S. adult population is estimated to have hypertension (1). Our ML approach also enables ongoing PA risk assessment updated from the EHR rather than a single blood-based snapshot assessment which may be confounded by medications and hypokalemia (6).

This ML model could enable a multi-phase screening approach by selectively prescreening patients with hypertension before standard screening assessment with serum/plasma aldosterone and renin. Aldosterone-renin ratio has >96% sensitivity and 52-61% specificity depending on cut-off threshold and costs $95 USD at Quest Diagnostics© as of August 2026 (28,29). A strength of the ML approach using EHR data is automated integration into clinical workflow, therefore improving screening rates. A quality improvement study showed that adding an alert to the EHR for PA testing improved screening rates (30). However, addressing low screening rates is one of the many barriers to successful implementation as challenges with interpretation of diagnostic work-up can also arise e.g. equivocal test results and special considerations for unilateral versus bilateral hyperaldosteronism demonstrated. These downstream effects have significant implications on health care delivery and endocrinology referrals. If current guidelines are strictly followed, it is estimated there will be an 800-fold increase in adrenalectomies in the United States per year (26). This is where AI solutions can potentially assist in triage, sequencing, interpretation, and clinical decision support to resolve such issues in healthcare delivery.

There are challenging technical aspects to developing an ML model for PA screening. The true prevalence of PA is unknown, and the majority of at-risk patients have not received screening or diagnostic testing. Therefore, it was necessary that the control cohort of the training dataset used patients who had negative screening tests, as the general population of patients with hypertension likely has undiagnosed PA among the unscreened population. However, selection bias may play a role as the training cohort draws from only patients who were identified to receive screening by a provider. Ultimately, the intent of this ML approach is not to diagnose PA but rather to improve screening methods with a targeted prescreening approach. The design and development of the model opted for a “supervised” approach to maximize transparency and explainability. We limited predictor variables to well-described features to mitigate healthcare access bias. Patients with increased healthcare access have more visit encounters and diagnoses, which may bias model predictions if all ICD code information were used. Obermeyer et al demonstrated how an ML model designed to triage sicker patients for higher-level care delayed referral along racial and socioeconomic lines due to this information bias (31). Therefore, we opted to design a model with predictors that would be less susceptible to high utilization and non-pertinent ICD codes, lab measurements, and medications outside those of interest in this present study.

Limitations exist within this study. This study draws predominantly from one institution, although the breadth spans geographically different regions and practice settings ranging from community clinics to tertiary/quaternary hospitals. The results may not be generalizable to other centers with different patient populations. The training dataset is limited by an incomplete ground truth as the true prevalence of PA is unknown and underdiagnosed. However, strengths include the large number of positive PA cases and patients who underwent screening. The simulation scenario exploring how our model would perform in a large cohort of patients with hypertension provides insight into real-world benefit. Future considerations of this work include model refinement, projected cost savings, and downstream quality/process improvement in referral patterns.

In conclusion, this ML model leverages routinely available EHR data to identify patients at increased risk for PA up to 1 year before screening or diagnosis. The model demonstrates acceptable discrimination, good calibration, and high specificity, supporting its potential role as a pragmatic prescreening tool to prioritize patients for guideline-directed biochemical testing. By utilizing explainable and commonly available clinical variables, this approach offers a scalable strategy for population-level risk stratification and may facilitate broader implementation of primary aldosteronism screening recommendations. Future external and prospective validation studies are needed to establish generalizability, clinical effectiveness, and impact on patient outcomes.

## Data Availability

All data produced are available online at mcpdiscover.org.

https://mcpdiscover.org/

## Funding Information

The Mayo Clinic Presidential Strategic Initiative Fund provided grant funding for associated model development and computing costs.

## Conflict of Interest and Disclosures

The authors acknowledge funding from the Mayo Clinic Presidential Strategic Initiative Fund. The authors have no relevant conflicts of interests. Chris Varghese has an unrelated relationship with Alimetry Ltd. Mayo Clinic received funding on behalf of William Perry for educational activities from Axonics (now Boston Scientific) and Cook Biotech (now Evergen), unrelated to this work. Wiliam Perry is an inventor of Mayo Clinic intellectual property which is licensed to HoloMedX LLC and may receive royalties paid to Mayo Clinic. Mayo Clinic received funding on behalf of Irina Bancos for consultation fees from AstraZeneca unrelated to this work. Mayo Clinic received research support on behalf of Irina Bancos from Esteve and Recordatti unrelated to this work. Cornelius Thiels received grant funding from Mayo Clinic Center for Digital Health Dalio Foundation AI/ML Enablement Award, Mayo Clinic Simons Family Career Development Award in Surgical Innovation, and Johnson and Johnson Polyphonic AI Fund for Surgery QuickFire Challenge unrelated to this work. Cornelius Thiels is an inventor of Mayo Clinic intellectual property which is licensed to apoQlar Medical and HoloMedX LLC and may receive royalties paid to Mayo Clinic. Cornelius Thiels served on the advisory board for Boston Scientific and Johnson & Johnson. All other authors have no conflicts of interest or disclosure to declare.

## Acknowledgments

Authors’ Contributions: Frank Lee wrote the initial draft of the manuscript. Ashok Choudhary and Leo Li-Han wrote the code for model development. Chris Varghese and Ellen Larson assisted with data curation and model conception. Irina Bancos, Trenton Foster, and Travis McKenzie assisted with interpretation of findings and provided valuable expertise towards the intellectual content. William Perry, Hojjat Salehinejad, and Cornelius Thiels supervised and were responsible for conception of the study and its design. All authors reviewed and made substantial contributions to the manuscript. Frank Lee and Ashok Choudhary had direct access to the underlying data and verified the accuracy at the participant-level. All authors had full access to the data in the study and take responsibility for the integrity of the data and the accuracy of the data analysis.

## Data Availability

Restrictions apply to the availability of some or all data generated or analyzed during this study to preserve patient confidentiality or because they were used under license. The corresponding author will on request detail the restrictions and any conditions under which access to some data may be provided.

## Supplementary Material

### S1: Mayo Clinic Platform data use statement

This study involves analysis of de-identified data via the Mayo Clinic Platform_Discover. In accordance with the Code of Federal Regulations, 45 CFR 46.102, the noted activity does not require IRB review. Data shown and reported in this manuscript has been extracted from this environment using an established protocol for data extraction, aimed at preserving patient privacy. The data has been de-identified pursuant to an expert determination in accordance with the HIPAA Privacy Rule. Any data beyond what is reported in the manuscript, including but not limited to the raw EHR data, cannot be shared or released due to the parameters of the expert determination to maintain the data de-identification

### S2: Inclusion criteria and schema tables for Mayo Clinic Platform cohort

| Data Type | Associated schema table |
| --- | --- |
| ICD codes | DIAGNOSIS_DIM_DIAGNOSIS_CODE/DIAGNOSIS_CODE |
| Labs | FACT_LAB_TEST/LAB_SUBTYPE_CODE |

| Primary aldosteronism-related ICD codes |
| --- |
| ICD-10 |
| E26.01 Conn's syndrome |
| E26.02 Glucocorticoid-remediable aldosteronism |
| E26.09 Other primary hyperaldosteronism |
| E26.89 Other hyperaldosteronism |
| E26.9 Hyperaldosteronism, unspecified |
| ICD-9 |
| 255.10 Hyperaldosteronism, unspecified |
| 255.12 Glucocorticoid-remediable aldosteronism |
| 255.12 Conn's syndrome |
| Hypertension-related ICD codes |
| ICD-10 |
| I10 Essential (primary) hypertension |
| I11.0 Hypertensive heart disease with heart failure |
| I11.9 Hypertensive heart disease without heart failure |
| I12.0 Hypertensive chronic kidney disease with stage 5 chronic kidney disease or end stage renal disease |
| I12.9 Hypertensive chronic kidney disease with stage 1 through stage 4 chronic kidney disease, or unspecified chronic kidney disease |
| I13.0 Hypertensive heart and chronic kidney disease with heart failure and stage 1 through stage 4 chronic kidney disease, or unspecified chronic kidney disease |
| I13.10 Hypertensive heart and chronic kidney disease without heart failure, with stage 1 through stage 4 chronic kidney disease, or unspecified chronic kidney disease |
| I13.11 Hypertensive heart and chronic kidney disease without heart failure, with stage 5 chronic kidney disease, or end stage renal disease |
| I13.2 Hypertensive heart and chronic kidney disease with heart failure and with stage 5 chronic kidney disease, or end stage renal disease |
| I15.0 Renovascular hypertension |
| I15.1 Hypertension secondary to other renal disorders |
| I15.2 Hypertension secondary to endocrine disorders |
| I15.8 Other secondary hypertension |
| I15.9 Secondary hypertension, unspecified |
| I16.0 Hypertensive urgency |
| I16.1 Hypertensive emergency |
| I16.9 Hypertensive crisis, unspecified |
| R03.0 Elevated blood-pressure reading, without diagnosis of hypertension |
| ICD-9 |
| 401.0 Malignant essential hypertension |
| 401.1 Benign essential hypertension |
| 401.9 Unspecified essential hypertension |
| 402.00 Malignant hypertensive heart disease without heart failure |
| 402.01 Malignant hypertensive heart disease with heart failure |
| 402.10 Benign hypertensive heart disease without heart failure |
| 402.11 Benign hypertensive heart disease with heart failure |
| 402.90 Unspecified hypertensive heart disease without heart failure |
| 402.91 Unspecified hypertensive heart disease with heart failure |
| 403.00 Hypertensive chronic kidney disease, malignant, with chronic kidney disease stage I through stage IV, or unspecified |
| 403.01 Hypertensive chronic kidney disease, malignant, with chronic kidney disease stage V or end stage renal disease |
| 403.10 Hypertensive chronic kidney disease, benign, with chronic kidney disease stage I through stage IV, or unspecified |
| 403.11 Hypertensive chronic kidney disease, benign, with chronic kidney disease stage V or end stage renal disease |
| 403.90 Hypertensive chronic kidney disease, unspecified, with chronic kidney disease stage I through stage IV, or unspecified |
| 403.91 Hypertensive chronic kidney disease, unspecified, with chronic kidney disease stage V or end stage renal disease |
| 404.00 Hypertensive heart and chronic kidney disease, malignant, without heart failure and with chronic kidney disease stage I through stage IV, or unspecified |
| 404.01 Hypertensive heart and chronic kidney disease, malignant, with heart failure and with chronic kidney disease stage I through stage IV, or unspecified |
| 404.02 Hypertensive heart and chronic kidney disease, malignant, without heart failure and with chronic kidney disease stage V or end stage renal disease |
| 404.03 Hypertensive heart and chronic kidney disease, malignant, with heart failure and with chronic kidney disease stage V or end stage renal disease |
| 404.10 Hypertensive heart and chronic kidney disease, benign, without heart failure and with chronic kidney disease stage I through stage IV, or unspecified |
| 404.11 Hypertensive heart and chronic kidney disease, benign, with heart failure and with chronic kidney disease stage I through stage IV, or unspecified |
| 404.12 Hypertensive heart and chronic kidney disease, benign, without heart failure and with chronic kidney disease stage V or end stage renal disease |
| 404.13 Hypertensive heart and chronic kidney disease, benign, with heart failure and chronic kidney disease stage V or end stage renal disease |
| 404.90 Hypertensive heart and chronic kidney disease, unspecified, without heart failure and with chronic kidney disease stage I through stage IV, or unspecified |
| 404.91 Hypertensive heart and chronic kidney disease, unspecified, with heart failure and with chronic kidney disease stage I through stage IV, or unspecified |
| 404.92 Hypertensive heart and chronic kidney disease, unspecified, without heart failure and with chronic kidney disease stage V or end stage renal disease |
| 404.93 Hypertensive heart and chronic kidney disease, unspecified, with heart failure and chronic kidney disease stage V or end stage renal disease |
| 405.01 Malignant renovascular hypertension |
| 405.09 Other malignant secondary hypertension |
| 405.11 Benign renovascular hypertension |
| 405.19 Other benign secondary hypertension |
| 405.91 Unspecified renovascular hypertension |
| 405.99 Other unspecified secondary hypertension |
| 796.2 Elevated blood pressure reading without diagnosis of hypertension |
| V81.1 Screening for hypertension |
| **Primary aldosteronism screening test name labels** |
| Aldosterone, P |
| Aldosterone, S |
| Aldosterone |
| Renin Activity, P |
| Renin Act |
| Renin Activity Plasma |
| Renin Activity-Mayo |

### S3: Model architecture algorithms used and their summary description

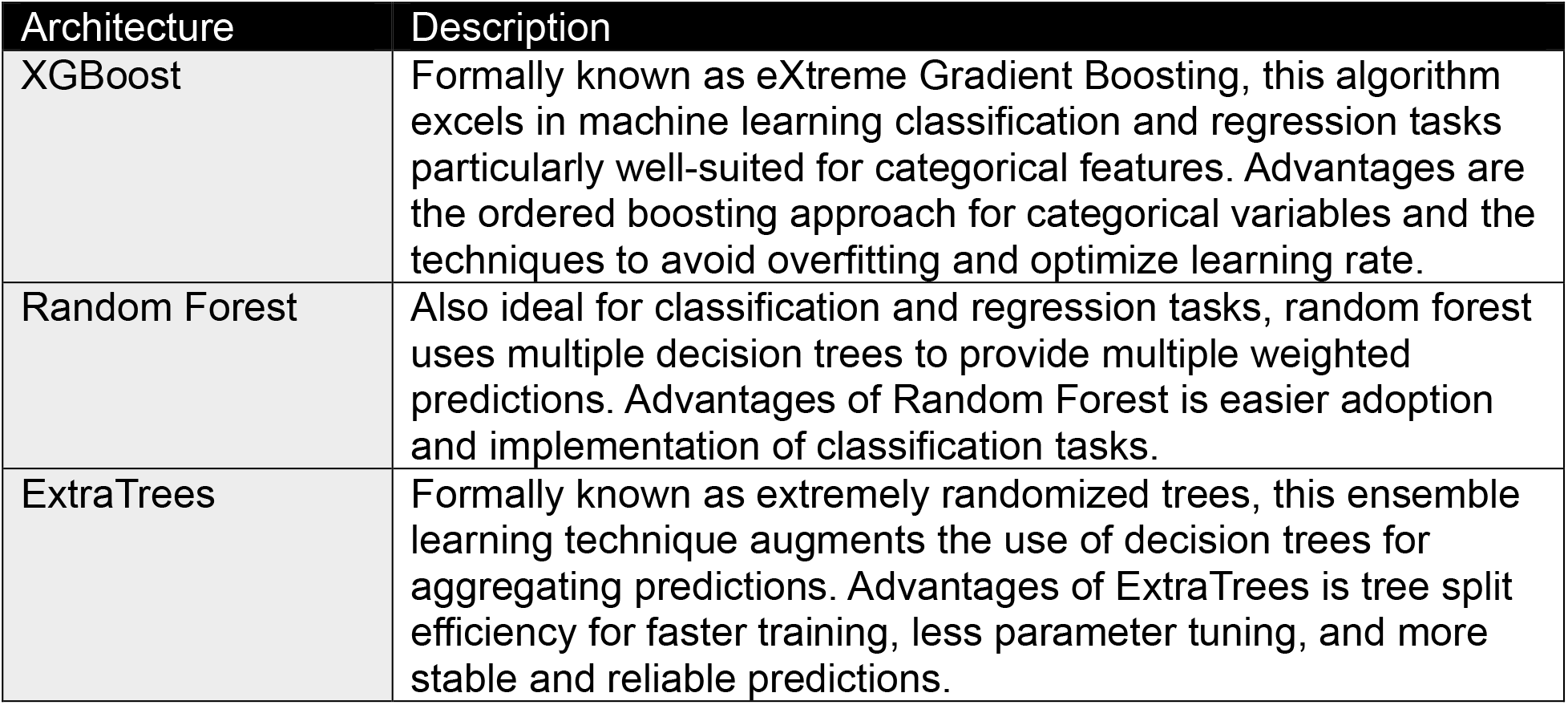

### S4: Predictor variable codes and associated schema tables in Mayo Clinic Platform

| Data Type | Associated schema table |
| --- | --- |
| ICD codes | DIAGNOSIS_DIM_DIAGNOSIS_CODE/DIAGNOSIS_CODE |
| Labs | FACT_LAB_TEST/LAB_SUBTYPE_CODE |
| Blood pressure | FACT_FLOWSHEETS/FLOWSHEET_SUBTYPE_DESCRIPTION |
| Medication | FACT_ORDERS/MED_GENERIC |
| Age | NFER_AGE |
| Sex | DIM_PATIENT/PATIENT_GENDER_NAME |

| Predictor ICD codes |
| --- |
| ICD-10 |
| E87.6 Hypokalemia |
| I10 Essential (primary) hypertension |
| I11.9 Hypertensive heart disease without heart failure |
| I12.9 Hypertensive chronic kidney disease with stage 1 through stage 4 chronic kidney disease, or unspecified chronic kidney disease |
| I15.8 Other secondary hypertension |
| I15.9 Secondary hypertension, unspecified |
| ICD-9 |
| 401.0 Malignant essential hypertension |
| 401.1 Benign essential hypertension |
| 401.9 Unspecified essential hypertension |
| 402.10 Benign hypertensive heart disease without heart failure |
| 403.10 Hypertensive chronic kidney disease, benign, with chronic kidney disease stage I through stage IV, or unspecified |
| 403.11 Hypertensive chronic kidney disease, benign, with chronic kidney disease stage V or end stage renal disease |
| 405.19 Other benign secondary hypertension |
| 405.99 Other unspecified secondary hypertension |
| 796.2 Elevated blood pressure reading without diagnosis of hypertension |
| Predictor labs* |
| Potassium, S |
| Potassium, P |
| K |
| Potassium Lvl |
| EXT Potassium, S |
| Potassium, B |
| EXT Potassium |
| Potassium, POCT, B |
| EXT Potassium, B |
| Potassium |
| Potassium Level |
| POC Potassium |
| EXT Potassium, POCT, B |
| Potassium, Venous, B |
| **Predictor vitals** |
| Systolic BP (mmHg) |
| Systolic Blood Pressure |
| SYSTOLIC BLOOD PRESSURE |
| **Predictor medications** |
| CHLORTHALIDONE |
| CHLOROTHIAZIDE |
| HYDROCHLOROTHIAZIDE |
| INDAPAMIDE |
| METOLAZONE |
| POLYTHIAZIDE |
| BUMETANIDE |
| FUROSEMIDE |
| TORSEMIDE |
| ATENOLOL |
| BETAXOLOL |
| BISOPROLOL |
| METOPROLOL |
| NADOLOL |
| PROPRANOLOL |
| PROPRANOLOL |
| TIMOLOL |
| ACEBUTOLOL |
| PENBUTOLOL |
| PINDOLOL |
| CARVEDILOL |
| LABETALOL |
| BENAZEPRIL |
| CAPTOPRIL |
| ENALAPRIL |
| FOSINOPRIL |
| LISINOPRIL |
| MOEXIPRIL |
| PERINDOPRIL |
| QUINAPRIL |
| RAMAPRIL |
| TRANDOLAPRIL |
| AZILSARTAN |
| CANDESARTAN |
| EPROSARTAN |
| IRBESARTAN |
| LOSARTAN |
| OLMESARTAN |
| TELMISARTAN |
| VALSARTAN |
| AMLODIPINE |
| FELODIPINE |
| ISRADIPINE |
| NICARDIPINE |
| NIFEDIPINE |
| NISOLDIPINE |
| DILTIAZEM |
| VERAPAMIL |
| AMILORIDE |
| TRIAMTERENE |
| EPLERENONE |
| SPIRONOLACTONE |
| POTASSIUM CHLORIDE |
| POTASSIUM CITRATE |

\*Any potassium lab measurement >10 was excluded due to likelihood of urinary (not blood or serum) sample

### S5: XGBoost Hyperparameter Search

| Hyperparameter | Setting |
| --- | --- |
| Number of estimators | 3200 |
| Learning rate | 0.01 |
| Maximum depth | 3 |
| Minimum child weight | 50 |
| Gamma | 5 |
| Subsample | 0.8 |
| Column sample by tree | 0.8 |
| Column sample by level | 0.8 |
| Column sample by node | 0.8 |
| L1 regularization (reg_alpha) | 100 |
| L2 regularization (reg_lambda) | 20 |
| Maximum delta step | 1 |

Random Forest Hyperparameters
| Hyperparameter | Setting |
| --- | --- |
| Number of estimators | 600 |

ExtraTrees Hyperparameters
| Hyperparameter | Setting |
| --- | --- |
| Number of estimators | 600 |

### S6: Formula definitions for machine learning model metrics

Sensitivity (a.k.a. Recall):

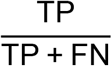

Specificity:

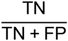

False Negative Rate (a.k.a. Miss Rate):

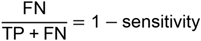

False Positive Rate:

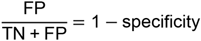

Accuracy:

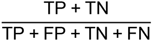

Precision (a.k.a. Positive Predictive Value):

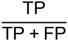

F1-score:

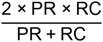

BS Brier score, TP true positive, TN true negative, FP false positive, FN false negative, PR precision, RC recall

### S7: Degree of missingness among input predictor variables

| Input predictor variable | Overall<br>(N = 22,264) | Disease cohort<br>(N = 1,833) | Control cohort<br>(N = 20,431) | *P* |
| --- | --- | --- | --- | --- |
| Missing ICD code (from inclusion criteria list) | 6,001 (27.0%) |  |  |  |
|  |  | 397 (21.7%) | 5,604 (27.4%) | <0.001<sup>x</sup> |
| Missing potassium lab | 8,714 (39.1%) |  |  |  |
|  |  | 873 (47.6%) | 7,841 (38.4%) | <0.001<sup>x</sup> |
| Missing medication (i.e. no antihypertensive or potassium supplementation prescription) | 15,787 (70.9%) |  |  |  |
|  |  | 1,331 (72.6%) | 14,456 (70.8%) | 0.099<sup>x</sup> |
| Missing blood pressure measurement | 8,884 (39.9%) |  |  |  |
|  |  | 765 (41.7%) | 8,119 (39.7%) | 0.100<sup>x</sup> |

Values reported as N (%); <sup>x</sup>Chi-square test

### S8: Equity analysis on race to assess model fairness across different subgroups

| Race | Total count | PA positive cases | Percent positive cases (within racial group) | AUROC | *P* |
| --- | --- | --- | --- | --- | --- |
| White/Caucasian | 19,831 | 1569 | 7.9% | 0.771 | 0.581 |
| Black/African American | 1,132 | 159 | 14.0% | 0.762 |  |
| Asian | 582 | 41 | 7.0% | 0.712 |  |
| Other | 719 | 64 | 8.9% | 0.747 |  |

Other includes Native American and Pacific Islander, undisclosed, and unknown

## Notes

### Author Declarations

Ethics committee/IRB of Mayo Clinic Rochester, Minnesota gave ethical approval for this work.

## References

1. Fryar CD, Kit B, Carroll MD, Afful J. Hypertension prevalence, awareness, treatment, and control among adults age 18 and older: United States, August 2021-Augst 2023. NCHS Data Brief, no 511. Hyattsville, MD: National Center for Health Statistics. 2024. doi:10.15620/cdc/164016

2. United States Census Bureau. National Population by Characteristics: 2020-2025. Revised April 2026. Accessed June 14, 2026. https://www.census.gov/data/tables/time-series/demo/popest/2020s-national-detail.html

3. Farah MH, Hegazi M, Firwana M, Abusalih M, Saadi S, Al-Kordi M, Elsheikh A, Wang Z, Hassett L, Bancos I, Murad MH. A systematic review supporting the Endocrine Society Clinical Practice Guideline on management of primary aldosteronism. J Clin Endocrinol Metab. 2025;110(9):e2833–e2844. doi:10.1210/clinem/dgaf290

4. Owei L, Wachtel H, Cohen JB. Broadening primary aldosteronism screening: alignment across contemporary guidelines. Hypertension. 2026;83(5):e26228. doi:10.1161/HYPERTENSIONAHA.125.26228

5. Adler GK, Stowasser M, Correa RR, Khan N, Kline G, McGowan MJ, Mulatero P, Murad MH, Touyz RM, Vaidya A, Williams TA, Yang J, Young WF, Zennaro MC, Brito JP. Primary aldosteronism: an Endocrine Society Clinical Practice Guideline. J Clin Endocrinol Metab. 2025;110(9):2453–2495. doi:10.1210/clinem/dgaf284

6. Reincke M, Bancos I, Mulatero P, Scholl UI, Stowasser M, Williams TA. Diagnosis and treatment of primary aldosteronism. Lancet Diabetes Endocrinol. 2021;9(12):876–892. doi:10.1016/S2213-8587(21)00210-2

7. Rossi GP, Rossi FB, Cignarella A, Seccia TM. Aldosterone synthase inhibitors for the treatment of cardiovascular disease. Nat Rev Endocrinol. 2026;22(9):543–553. doi:10.1038/s41574-026-01259-4

8. Carling T, Faucz FR. Primary aldosteronism: adrenalectomy could save more lives. Front Endocrinol. 2025;16:1737160. doi:10.3389/fendo.2025.1737160

9. Ruhle B, Livhits, MJ. Routine versus selective screening for primary aldosteronism in patients with hypertension. In: James BC, Grogan RH, Angelos P, eds. Difficult Decisions in Endocrine Surgery: An Evidence-Based Approach. 2nd ed. Springer; 2025, 331–344.

10. Williams JS, Williams GH, Raji A, Jeunemaitre X, Brown NJ, Hopkins PN, Conlin PR. Prevalence of primary hyperaldosteronism in mild to moderate hypertension without hypokalaemia. J Hum Hypertens. 2006;20(2):129–136. doi:10.1038/sj.jhh.1001948

11. Placido D, Yuan B, Hjaltelin JX, Zheng C, Haue AD, Chmura PJ, Yuan C, Kim J, Umeton R, Antell G, Chowdhury A, Franz A, Brais L, Andrews E, Marks DS, Regev A, Ayandeh S, Brophy MT, Do NV, Kraft P, Wolpin BM, Rosenthal MH, Fillmore NR, Brunak S, Sander C. A deep learning algorithm to predict risk of pancreatic cancer from disease trajectories. Nat Med. 2023;29(5):1113–1122. doi:10.1038/s41591-023-02332-5

12. Mukherjee P, Humbert-Droz M, Chen JH, Gevaert O. SCOPE: predicting future diagnoses in office visits using electronic health records. Sci Rep. 2023;13(1):11005. doi:10.1038/s41598-023-38257-9

13. Mavromatis LA, Zlatanic V, Agarunov E, Sanoba SA, Kluger MD, Horwitz LI, Razavian N, Maitra A, Gonda TA, Grams ME. Development and validation of a parsimonious risk stratification model for pancreatic cancer. JAMA Oncol. Available online ahead of print March 26, 2026. doi:10.1001/jamaoncol.2026.0372

14. Wang HH, Wang YH, Liang CW, Li YC. Assessment of deep learning using nonimaging information and sequential medical records to develop a prediction model for nonmelanoma skin cancer. JAMA Dermatol. 2019;155(11):1277–1283. doi:10.1001/jamadermatol.2019.2335

15. Moons KGM, Damen JAA, Kaul T, Hooft L, Andaur Navarro C, Dhiman P, Beam AL, Van Calster B, Celi LA, Denaxas S, Denniston AK, Ghassemi M, Heinze G, Kengne AP, Maier-Hein L, Liu X, Logullo P, McCradden MD, Liu N, Oakden-Rayner L, Singh K, Ting DS, Wynants L, Yang B, Reitsma JB, Riley RD, Collins GS, van Smeden M. PROBAST+AI: an updated quality, risk of bias, and applicability assessment tool for prediction models using regression or artificial intelligence methods. BMJ. 2025;388:e082505. doi:10.1136/bmj-2024-082505

16. Yu Y, Hu X, Rajaganapathy S, Feng J, Abdelhameed A, Li X, Li J, Liu X, Yang L, Ertekin-Taner N, Fiero P, Boroumand S, Larsen R, Goyal M, Otley CC, Zong N, Shah VH, Halamka JD, Tao C. Accelerating AI innovation in healthcare: real-world clinical research applications on the Mayo Clinic Platform. Npj Health Syst. 2026;3(1):17. doi:10.1038/s44401-026-00068-1

17. Ganguly A. Primary aldosteronism. N Engl J Med. 1998;339(25):1828–1834. doi:10.1056/NEJM199812173392507

18. Akasaka H, Yamamoto K, Rakugi H, Nagasawa M, Nakamaru R, Ichijo T, Takeda Y, Kurihara I, Katabami T, Tsuiki M, Wada N, Ogawa Y, Kawashima J, Sone M, Kamemura K, Yoshimoto T, Matsuda Y, Fujita M, Kobayashi H, Watanabe M, Tamura K, Okamura S, Miyauchi S, Izawa S, Chiba Y, Tanabe A, Naruse M, Japan Primary Aldosteronism Study Group. Sex difference in the association between subtype distribution and age at diagnosis in patients with primary aldosteronism. Hypertension. 2019;74(2):368–374. doi:10.1161/HYPERTENSIONAHA.119.13006

19. Hosmer DW, Lemeshow S, Sturdivant RX. Applied Logistic Regression. Hoboken: Wiley; 2013; 176–177.

20. Libianto R, Fuller PJ, Young MJ, Yang J. Primary aldosteronism is a public health issue: challenges and opportunities. J Hum Hypertens. 2020;34(7):478–486. doi:10.1038/s41371-020-0336-2

21. Mulatero P, Monticone S, Burrello J, Veglio F, Williams TA, Funder J. Guidelines for primary aldosteronism: uptake by primary care physicians in Europe. J Hypertens. 2016;34(11):2253–2257. doi:10.1097/HJH.0000000000001088

22. Cohen JB, Cohen DL, Herman DS, Leppert JT, Byrd JB, Bhalla V. Testing for primary aldosteronism and mineralocorticoid receptor antagonist use among U.S. veterans: a retrospective cohort study. Ann Intern Med. 2021;174(3):289–297. doi:10.7326/M20-4873

23. Gupta G, Teo AED, Swee DS, Loh LM, Chuah M, Loh WJ, Saffari SE, Koh XH, Kek PC, Puar TH. Prolonged hypokalemia and delayed diagnosis of primary aldosteronism: clinical course and risk factors. J Clin Endocrinol Metab. 2024;109(8):e1574–e1581. doi:10.1210/clinem/dgad752

24. McEvoy JW, McCarthy CP, Bruno RM, Brouwers S, Canavan MD, Ceconi C, Christodorescu RM, Daskalopoulou SS, Ferro CJ, Gerdts E, Hanssen H, Harris J, Lauder L, McManus RJ, Molloy GJ, Rahimi K, Regitz-Zagrosek V, Rossi GP, Sandset EC, Scheenaerts B, Staessen JA, Uchmanowicz I, Volterrani M, Touyz RM; ESC Scientific Document Group. 2024 ESC guidelines for the management of elevated blood pressure and hypertension. Eur Heart J. 2024;45(38):3912–4018. doi:10.1093/eurheartj/ehae178

25. Jones DW, Ferdinand KC, Taler SJ, Johnson HM, Shimbo D, Abdalla M, Altieri MM, Bansal N, Bello NA, Bress AP, Carter J, Cohen JB, Collins KJ, Commodore-Mensah Y, Davis LL, Egan B, Khan SS, Lloyd-Jones DM, Melnyk BM, Mistry EA, Ogunniyi MO, Schott SL, Smith SC Jr, Talbot AW, Vongpatanasin W, Watson KE, Whelton PK, Williamson JD. 2025 AHA/ACC/AANP/AAPA/ABC/ACCP/ACPM/AGS/AMA/ASPC/NMA/PCNA/SGIM guideline for the prevention, detection, evaluation and management of high blood pressure in adults: a report of the American College of Cardiology/American Heart Association Joint Committee on Clinical Practice Guidelines. Hypertension. 2025;82(10):e212-e316. doi:10.1161/HYP.0000000000000249

26. Carling T, Patel K, Stratakis CA, Faucz FR. Tailored vein sampling and surgery in primary aldosteronism. JAMA Surg. 2026;161(5):443–445. doi:10.1001/jamasurg.2026.0157

27. Wilson JMG, Jungner G. Principles and Practice of Screening for Disease. World Health Organization; 1968.

28. Liu X, Hao S, Bian J, Lou Y, Zhang H, Wu H, Cai J, Ma W. Performance of aldosterone-to-renin ratio before washout of antihypertensive drugs in screening of primary aldosteronism. J Clin Endocrinol Metab. 2024;109(12):e2302–e2308. doi:10.1210/clinem/dgae094

29. Quest Diagnostics. Primary aldosteronism test. Quest. Accessed June 3, 2026. https://www.questhealth.com/product/primary-aldosteronism-test/13817M

30. Charoensri S, Bashaw L, Dehmlow C, Ellies T, Wyckoff J, Turcu AF. Evaluation of a best-practice advisory for primary aldosteronism screening. JAMA Intern Med. 2024;184(2):174–182. doi:10.1001/jamainternmed.2023.7389

31. Obermeyer Z, Powers B, Vogeli C, Mullainathan S. Dissecting racial bias in an algorithm used to manage the health of populations. Science. 2019;386:447–453.

